# Assessment of burden and risk factors associated with Soil-transmitted helminth infections among adolescent girls in Katete District of Zambia: A Cross-Sectional Study

**DOI:** 10.1101/2025.03.21.25324433

**Authors:** Buumba Tapisha, Westone P. Hamwata, Danny Sinyange, Hikabasa Halwindi, Adamson S. Muula

**Affiliations:** The University of Malawi, College of Medicine, Private Bag 360, Blantyre, Malawi, Africa; Tropical Diseases Research Centre, Biomedical Sciences Department, P.O Box 71769, Ndola, Zambia; Zambia National Public Health Institute, P.O Box ……., Lusaka, Zambia; The University of Zambia, School of Public Health Department of Public Health, P.O Box 50516, Lusaka, Zambia; The University of Malawi, College of Medicine, Department of Public Health, Private Bag 360, Blantyre Malawi

**Author notes:** Corresponding Author: Westone Hamwata.

**Keywords:** **Keyword**s: Soil Transmitted Helminths, risk factors, girls, Women of Reproductive Age, Katete District, Zambia

## Abstract

**Background:** Over 688 million girls and women of reproductive age (WRA) are at risk of Soil-Transmitted Helminths (STH) infections, with 26% of girls and WRA found in Africa. Infections among girls and WRA remain a concern because of their association with anaemia among non-pregnant girls and women, maternal anaemia, foetal morbidity, and mortality. Information regarding subpopulations such as adolescent girls in the district at risk remains unknown, making it difficult to plan and implement interventions as guided in the new NTD roadmap 2021-2030. This study assessed the burden and risk factors associated with Soil-transmitted helminth infections among adolescent girls in Katete District of Zambia.

**Methodology:** A cross-sectional study was conducted to gather data from 206 adolescent girls aged 10 to 19years between August and September, 2021. Multi-stage sampling was used to randomly select participants from 12 public schools. The study employed a structured questionnaire and analyzed stool samples using Kato-Katz to gather pertinent information on socio-demographic, socio-economic, behavioral, environmental health-related data, and prevalence respectively.. Additionally, logistic regression was used to identify factors associated with Soil-Transmitted Helminth Infections in the district.

**Results:** The prevalence of STH infections was 9.9% (19/192) and the only parasites found was *Ascaris lumbricoides.* The overall intensity was light infection. The factors independently associated with STH infection after adjusting for other variables were dirty fingernails (AOR: 0.2 95%CI 0.06-0.67, p=0.008) and drinking water directly from the source (AOR: 3.80 95%CI 1.19-12.15, p=0.024)

**Conclusion:** The findings indicate that STH infections are still prevalent. Interventions employed to further reduce the prevalence should include or strengthen behavioral change that addresses factors related to personal hygiene and drinking clean and safe water.

## Background

The call to end neglect of neglected tropical diseases (among others Soil-transmitted helminths STH) by WHO member states present an opportunity assist over 1.5 billion people infected worldwide, freeing them from “unnecessary suffering, stigma, and neglect”. The highest numbers of those infected occur in Sub-Saharan Africa, the Americas, China, and East Asia^1,2^. It is further estimated that over 688 million girls and women of reproductive age (WRA) are at risk of STH infections, with 26% of girls and WRA found in Africa ^2^. It is for this reason that the WHO has set an ambitious goal to end the Neglect of NTDs in an effort to attain the Sustainable Development Goals (SDGs) 2021-2030 through advocacy among other things, accelerating actions to reduce incidence, prevalence, morbidity, disability, and death in extreme events caused by STH.^1,2^

The most prevalent and widely distributed STHs species affecting people include roundworms (*Ascaris Lumbricoides*), hookworms (*Nector americanus and Ancylostoma),* and whipworms (*Trichuris trichiuria*). These species get addressed as a group because they are diagnosed and treated in a similar manner ^1^. STH infections are associated with reduced productivity, malnutrition, retarded growth, and retarded cognitive development ^3–12^. These infections reduce work capacity and impede concentration. In children, infections are associated with impaired nutrition status causing loss of iron and anaemia, reduced cognitive development, and increased absenteeism from school. Infections are also associated with maternal anaemia and foetal morbidity and mortality. These infections are prevalent in poverty-stricken communities with poor environmental hygiene, and impoverished health services ^1,13,14^.

The effects of STH on adolescent girls are of particular concern because of their association with increased blood loss (girls already lose blood during menstruation making this blood loss harmful) which increases the risks of iron-deficiency anaemia, infant mortality, and low birth weight^15^. The benefits of deworming adolescent girls (and women of reproductive age) include but are not limited to preventing the agglomeration of worms, reducing or protecting against anaemia (and its consequences), and generally better preparing for future pregnancy. The other benefit is that the risk of low birthweight and neonatal mortality as a result of STH is reduced^15,16^. WHO recommends that preschool children (PSAC), school-aged girls, non-pregnant adolescent girls, and women of reproductive age living in STH endemic areas be dewormed with albendazole or mebendazole. This category of people is classified as an at-risk population^15^. The global coverage of deworming for PSAC and school-aged children (SAC) between 2010 and 2019 showed a 29% increase from 31% to 60% (20). However, deworming coverage of adolescent girls and women of reproductive age has remained low^17,18^. According to the Bellagio Declaration, every girl has a right to be treated when infected with soil transmitted helminths^19,20^. This implies that coverage for this at-risk population needs to be increased.

Zambia is one of the countries endemic to Soil-transmitted helminths targeted for elimination through Mass Drug Administration (MDA) and case management. The country has had an active NTD control program since the year 2003 with seven different NTD’s targeted for control and elimination^21^. In the year 2012, 115 districts were known to be endemic to STHs with over 1.7 million people estimated to be infected. Cross-sectional surveys carried out between 2014 and 2019 in Luangwa, Chililabombwe, Kafue, Mazabuka, Kalabo, and Serenje reveal a prevalence between 2 and 35%.^22–24^ These were studies conducted on PSAC aged below 5 years and SAC aged 5-15 years. A study conducted in two districts in the southern province of Zambia among adults aged 18 years and older revealed a combined STH prevalence of 7.4% to 12.1% ^25^. This shows that adults can also be a potential source of infections, especially in areas with open defecation, poor hygiene and poor drinking water supply.

In 2020 alone, Katete district recorded a total of 659 cases of intestinal infections cases from its health facilities^26^. These cases could be attributed to among other factors open defecation, poor water, and sanitation coverage. The possibility of having many unreported cases in the community is high. Deworming programs in the district mostly target under 5-year-old children during the bi- annual child health week programs^16^. School-aged children (5-14 years) are dewormed in schools at least once a year but are subject to the availability of deworming drugs. A gap between the PSAC and adults needs to be understood in terms of burden in this age group. This study tries to fill that gap by investigating the burden and risk factors among adolescent girls (because of the consequences STH can have on their wellbeing). This study will contribute to the knowledge and understanding of the burden and risk factors of STH in Zambia among adolescent girls. Proper estimates of the disease burden among adolescent girls largely remain unknown.

## Methods

### Study design and setting

A school-based cross sectional study was conducted among adolescent girls aged between 10 and 19 in 12 schools of Katete District. The study was conducted from May 2021 to October 2021. This timeframe encompassed various stages of the study such as proposal preparation, submission for approval, approval process, training, pretesting of the questionnaire, data collection, data analysis, report preparation, and dissemination of the findings. Katete District (-11.2166658, 33.1499994) is situated in the Eastern Province of the Republic of Zambia and is one of the fourteen districts in the area.. The district lies along the Great East Road about 87 Kilometers South- West of the Provincial Capital, Chipata, and approximately 500 Kilometers away from Lusaka, the Capital City of the Country. It has a total of 141 schools including private and public schools^27^. The schools are usually used as health service points public health interventions that target school children such as deworming, Human Papillomavirus Vaccination (HPV) and many others. The primary outcome of interest was infection status defined as infected (having at least one of the three STH species/eggs isolated in stool sample) or not infected.

### Study population

The study population in this study comprised of all adolescent girls aged between 10 and 19 years enrolled in Katete District schools. Data collection was however conducted in the 12 schools that were sampled.

### Inclusion and exclusion criteria

Adolescent girls that were present in class and willing to obtain informed assent from their guardians were included in the study. Adolescent girls who refused to provide freshly collected stool samples were not included in the study.

### Sample size estimation

The sample size was estimated using a formula for survey sample size estimation at 95% level of confidence^16^.

Where; n

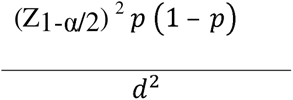

n = sample size, Z_1-α/2_= value of the standard normal distribution (1.96) corresponding to a significance level of 0.05 for a 2-sided test, d = margin of error (0.05), the prevalence (p= 0.1281) assumption of the study was based on a national survey conducted in Katete in 2012 ^28^. The sample size calculated was 172 participants. Adding a 20% to account for non-responses increased the sample size to 206 participants. This sample size was based on sample size determination for health studies when a survey is carried out in an ecologically homogenous area to evaluate prevalence and intensity which sets an adequate sample between 200 and 250 individuals ^29^

### Sampling strategies and data collection

Multistage sampling was used to select the study participants. The district has 10 zones that have a total of 141 private and public government schools. Cluster random sampling was used to sample 4 zones and a stratified random method to select 3 schools from each zone to come up with 12 schools. A systematic sampling method was used to select 17 study participants from each school through aggregated registers of girls in the age group 10-19 years (with two schools meant to contribute 18 participants each) to arrive at 206 study participants. The data used for identifying associated risk factors and burden of STH in WRA was collected using structured questionnaires and stool samples respectively.). To validate the questionnaire, a pilot study of 30 participants was conducted at Omelo Mumba School in a similar study setting away from the study site to optimize the questionnaire. After optimization of the data, the validated version of the data collection tools was employed in the study.

Stool samples were collected using tight and leak-proof containers. Each study participant was given a container for stool collection before the interview so that the sample was collected after the interview. The samples were collected at schools. The containers were labeled with the participant’s identification number, age, sex, time, and date of collection before giving the container to the participant. The samples were stored and transported in cool boxes at least within 1 hour of collection to the laboratory for analysis.

### Quality control

The questionnaires were checked for errors, completeness, or missing data at the end of each day. The data clerk completed the coding of all relevant fields on the questionnaire and entered them into an excel database. All the samples were submitted to the laboratory technician for analysis using the Kato-Katz technique within 3 hours of sample collection. Sample results were read by two independent technicians for quality assurance & results were entered into a secure database.

### Statistical Analysis

All statistical analyses were performed using STATA software version 14. Proportions for infected were computed to report the prevalence and intensity of infection. Crude egg count for soil-transmitted helminths was classified as light for eggs less than 5000EPG and moderate above 5000 to 49000 and heavy above 50000 when comparing egg count values between different ages, gender, and catchment areas^30^. Categorical variables were reported as absolute frequencies with associated percentages, and Chi-square tests were performed to ascertain the association between categorical independent variables and risk factors. Multivariate Logistic regression was used to assess factors independently associated with soil-transmitted helminths with a 95% confidence interval.

### Ethical Considerations

Ethics clearance to undertake this study was obtained from the Tropical Diseases Research Centre Ethics Committee (**Reference No. TRC/C4/05/2021**) and National Health Research Authority (**Ref No: NHRA000013/31/05/2021**). At District level, the permission to conduct this study in schools was sought from the Katete District Education Board Secretary (DEBS). Additionally, written assent was obtained from the parents or guardians of the participants prior to administration of the questionnaire and sample collection.

## Results

### Demographic Characteristics of Study Population

A total of 12 schools were surveyed out of 141 Schools in the district. A total of 192/206 (93%) pupils were interviewed and provided stool samples. Fourteen pupils did not submit stool samples and were not included in the analysis. The mean age of all participants was 14.26 years (S.D 2.24 years). The proportion of girls at the primary school level was significantly higher than those at the secondary school level (*p*<0.001). The summary of the characteristics is provided in table 1.

**Table 1:**
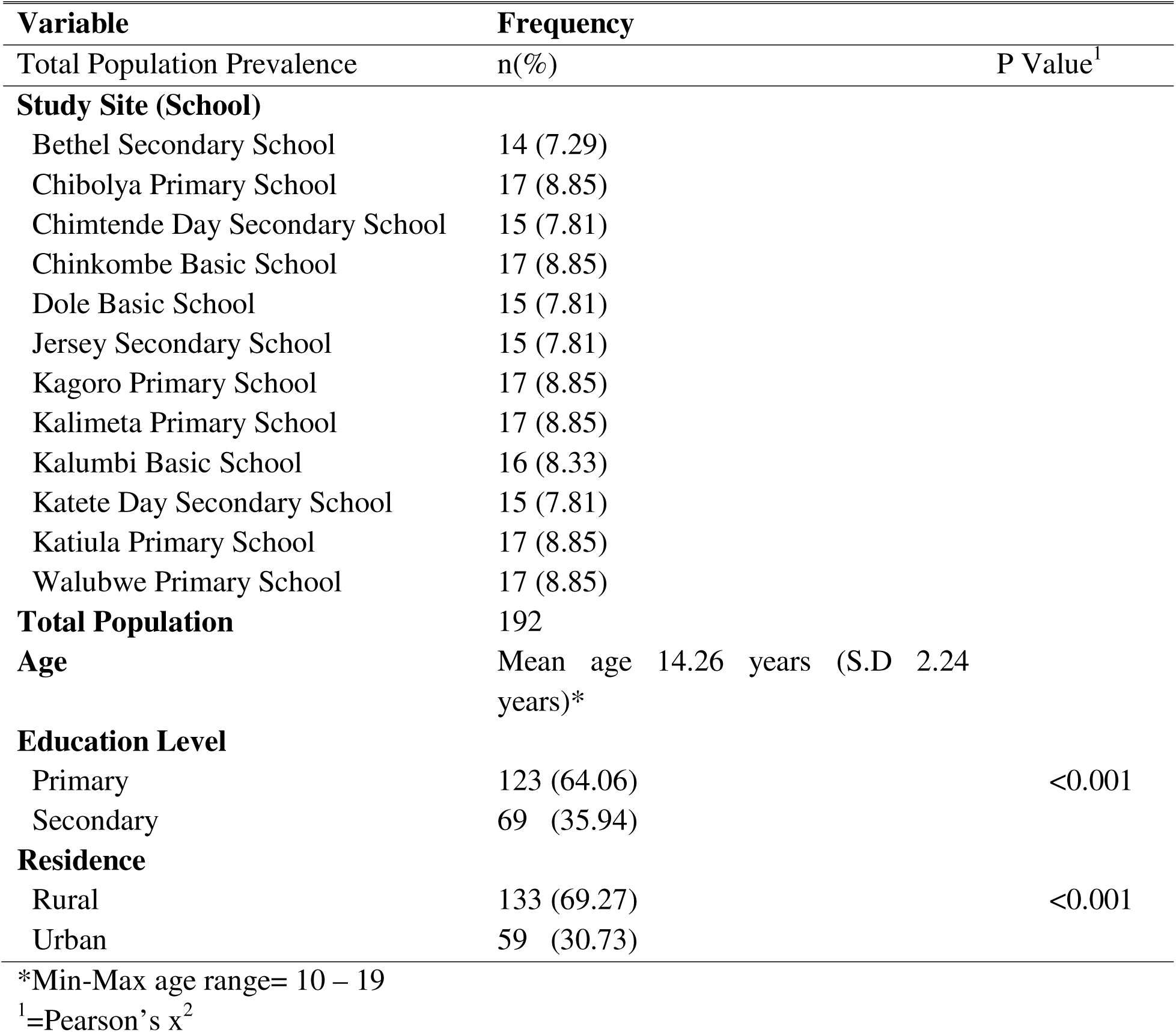
Characteristics of Study Population.

| **Variable** | **Frequency** |  |
| --- | --- | --- |
| Total Population Prevalence | n(%) | P Value<sup>1</sup> |
| **Study Site (School)** |  |  |
| Bethel Secondary School | 14 (7.29) |  |
| Chibolya Primary School | 17 (8.85) |  |
| Chimtende Day Secondary School | 15 (7.81) |  |
| Chinkombe Basic School | 17 (8.85) |  |
| Dole Basic School | 15 (7.81) |  |
| Jersey Secondary School | 15 (7.81) |  |
| Kagoro Primary School | 17 (8.85) |  |
| Kalimeta Primary School | 17 (8.85) |  |
| Kalumbi Basic School | 16 (8.33) |  |
| Katete Day Secondary School | 15 (7.81) |  |
| Katiula Primary School | 17 (8.85) |  |
| Walubwe Primary School | 17 (8.85) |  |
| **Total Population** | 192 |  |
| **Age** | Mean age 14.26 years (S.D 2.24 years)* |  |
| **Education Level** |  |  |
| Primary | 123 (64.06) | <0.001 |
| Secondary | 69 (35.94) |  |
| **Residence** |  |  |
| Rural | 133 (69.27) | <0.001 |
| Urban | 59 (30.73) |  |

\*Min-Max age range= 10 – 19
<sup>1</sup>=Pearson's $\chi^2$

### Prevalence and intensity

The prevalence of STH and infected population by schools are tabulated in fig 1. *Ascaris lumbricoides* was the only STH found in the samples analyzed from the 12 schools that participated in the study and gave an aggregated prevalence of 9.90% (19/192). The STH infection was highest in those attending primary school (grade one to seven) at 7.81% compared to those in secondary school level at 2.08%. The prevalence range was 0 to 25% across schools. The study revealed that 68.4% (13/19) of the pupils had light infection, 31.58% (6/19) of the pupils had moderate infections and no heavy infection was recorded. This infection is for the parasite found *A. lumbricoides*

**Fig 1.**
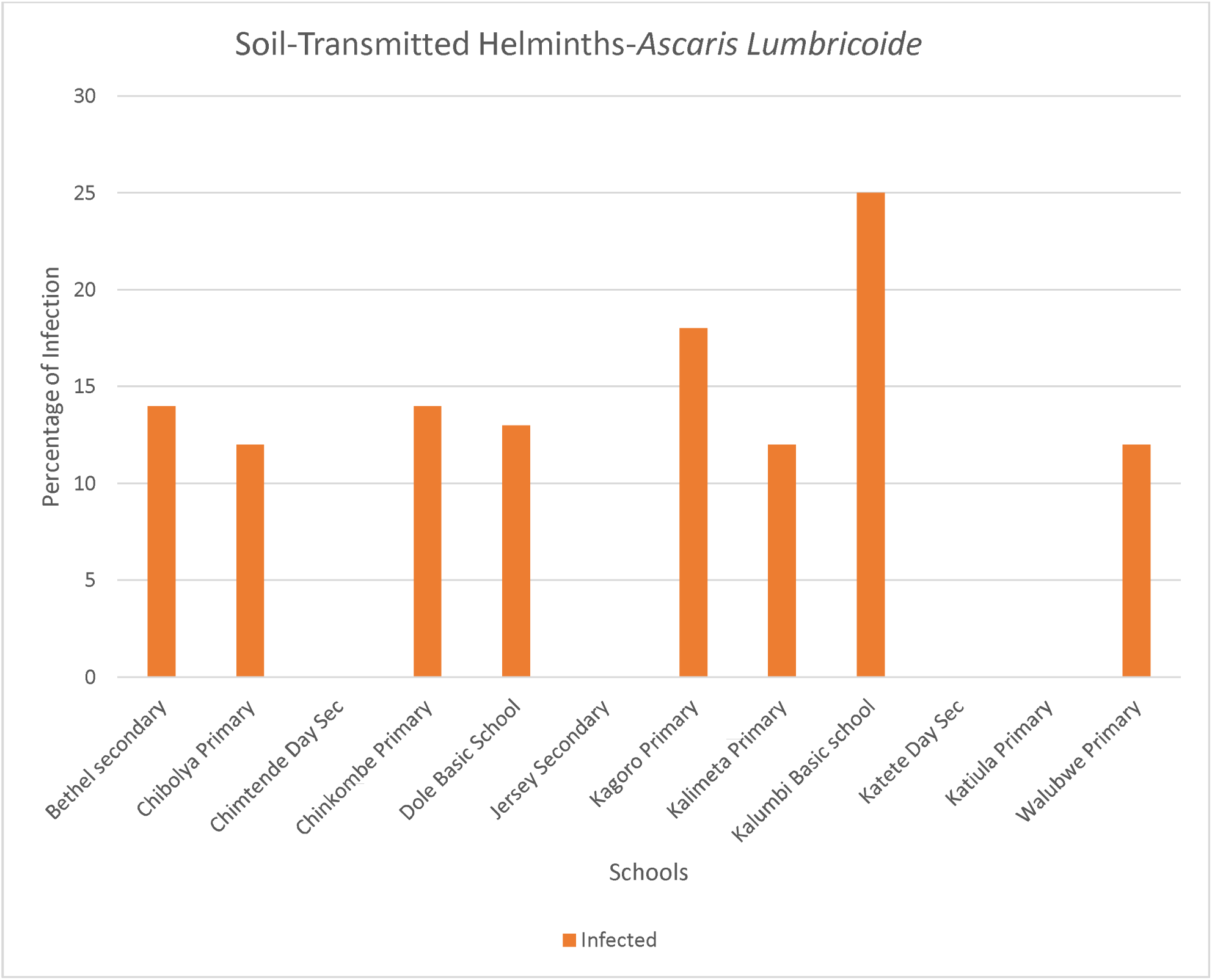
Prevalence of *Ascaris lumbricoides* in Schools.

### Risk factors Associated with STH Infections

#### Univariate Analysis

The participants in the age group 15-19 years were 78% less likely to be infected than participants in the age group 10-14years (OR: 0.22, 95% CI 0.06 – 0.78, p=0.019) as can be seen in table 2. The participants at the secondary school level were about half less likely to be infected than those at the primary school level although the difference was not statistically different (OR: 0.44, 95% CI 0.14 – 1.39, p=0.164). The study also showed that participants in urban areas had a 43% chance of not being infected with soil-transmitted helminths compared to those in rural areas. This difference was not statistically different (OR: 0.57, 95% CI 0.18 – 1.80, p=0.341). Participants whose parents or guardians were unemployed were 5 times more likely to be infected than those whose parents were formally employed. The difference observed was, however, not statistically significant (OR: 4.78, 95% CI 0.60 – 38.04, p=0.139).

**Table 2:**
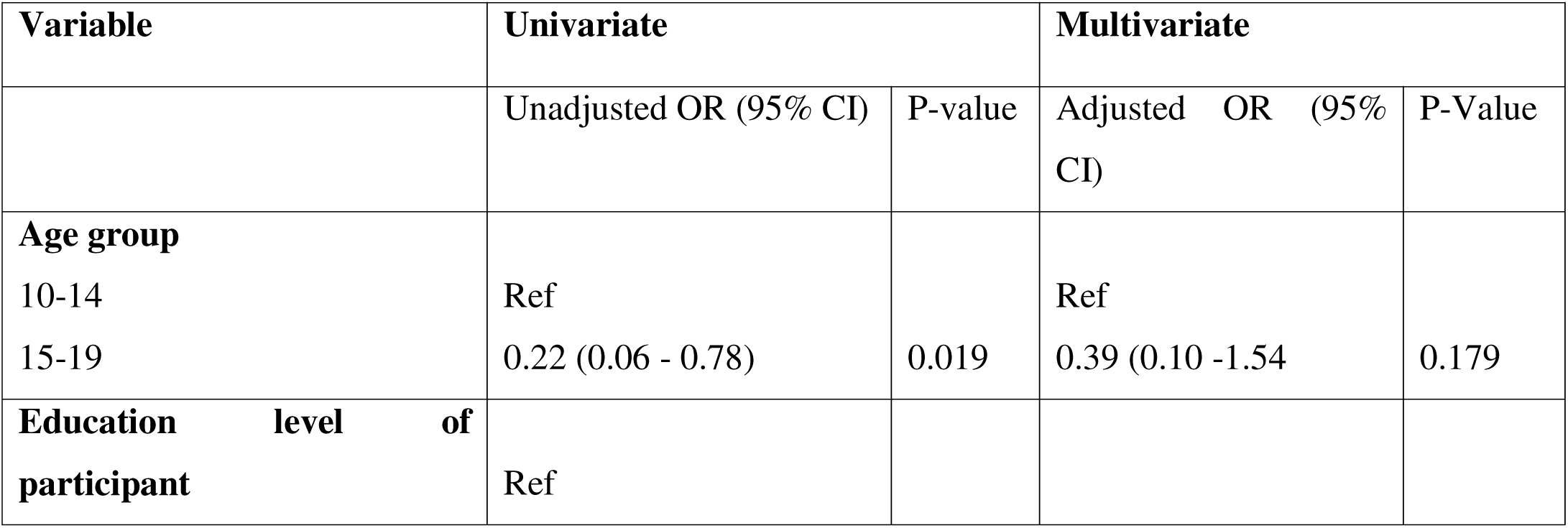
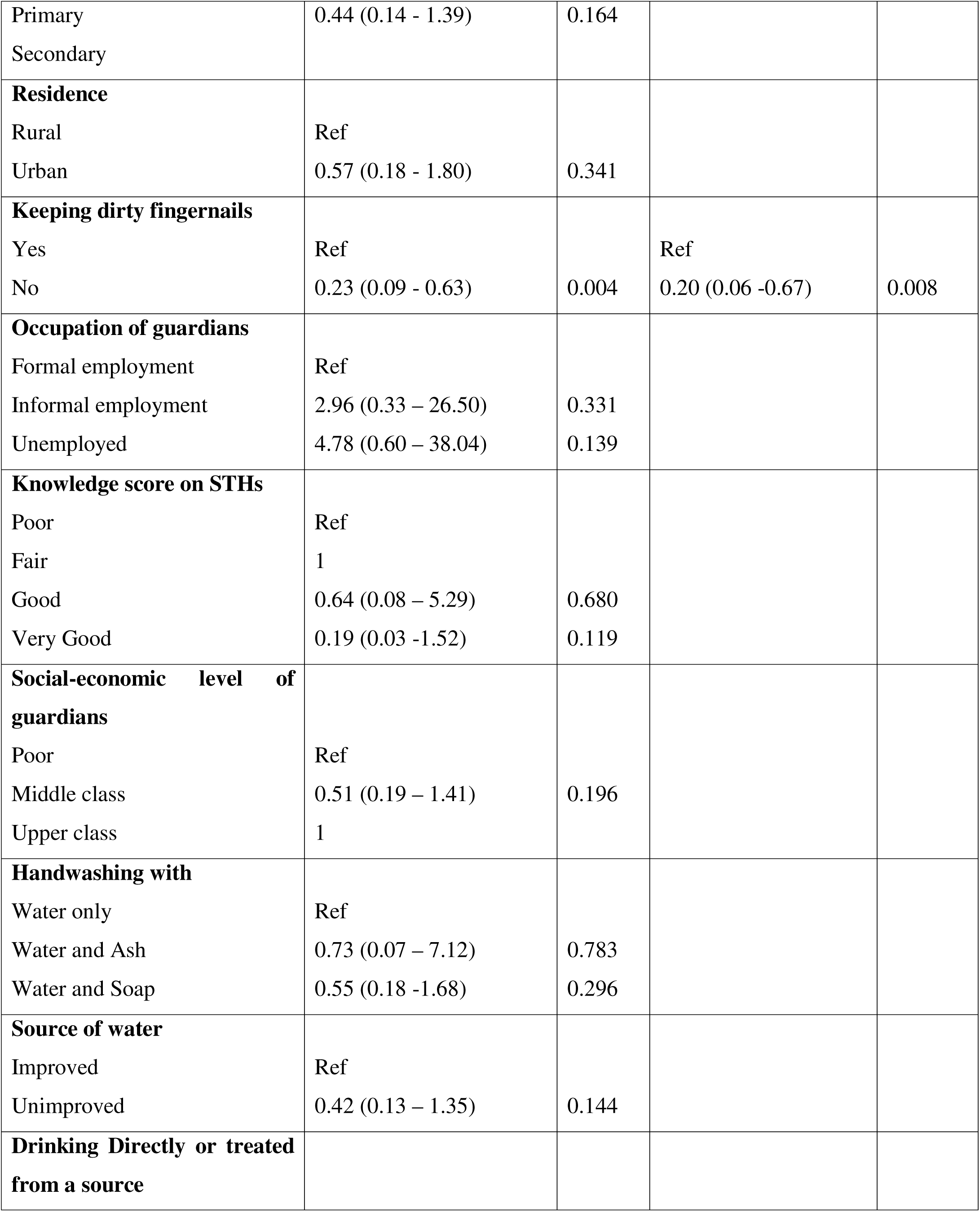
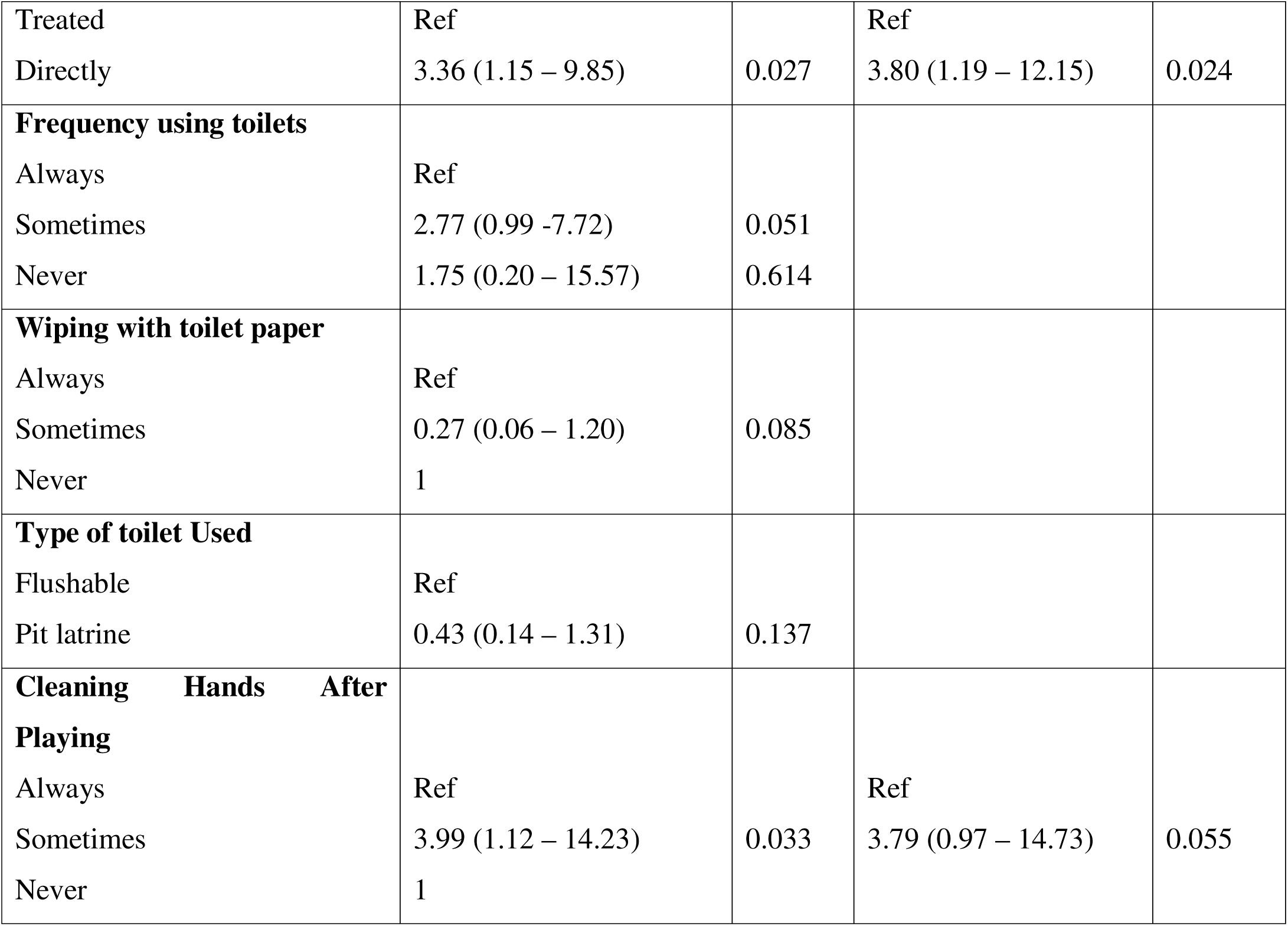
Univariate and Multivariate Analysis.

#### Multivariate Analysis

The variables with a p-value of less than 0.05 were further run in a multivariate analysis. Predictors of STH infections were dirty nails with a lower risk for those not keeping dirty fingernails (AOR: 0.2, 95%CI 0.06 – 0.67, p=0.008), and those drinking water directly from the source were 4 times more at risk (AOR:3.80, 95% CI 1.19 – 12.15, p=0.024), as shown in table 2.

## Discussion

This study sought to understand the burden and risk factors of STH among adolescent girls in Katete District. The study findings showed that *Ascaris lumbricoides* was the only STH found in this study with a prevalence of 9.90%. The prevalence of hookworms (*Nector americanus and Ancylostoma),* and whipworms (*Trichuris trichiuria* was found to be zero percent. *Ascaris lumbricoides* as seen in this study and earlier studies conducted, is the predominant and widely distributed STH in Zambia and other endemic countries ^22,25,31^. Despite the concerning intestinal infections recorded in Katete district, the prevalence of STH is lower than prevalence’s reported in Chililabombwe, Siavonga, Luangwa and Ndola. This is an indication that there may be other causes of intestinal infections in the district that warrants further investigation.

Most studies conducted on STH infections traditionally focused on SAC aged of 6 and 15, because the main public health intervention against STH infections is deworming which is school-based^25^. WHO intends to eliminate morbidity caused by STH infections in a previously neglected high-risk group of adolescent girls, pregnant and lactating mothers by the year 2030 ^15^. The results of this study suggest that if adolescent girls infected in schools are left untreated, they may perpetuate the transmission of STH infections in areas they come from thus negating the efforts being made to achieve the 2030 goal of eliminating or reducing the prevalence to less than 2%. This is a potential problem, especially in areas where MDA initiatives are not implemented or where only children under 5 years are targeted as is the case for Katete district.

Other studies conducted in Zambia have shown that STH infections are endemic with varying prevalence and intensity ^22–25^. The prevalence recorded in this study was lower than the prevalence in a survey conducted in 2012 by the Ministry of Health which was 12.81% ^21^. A study conducted in the Copperbelt Province of Zambia in school-aged children between 5 and 15 years found a prevalence of 14.4%.^22^.

The intensity of the infections was mostly light infections with few moderate infections, despite 97% not being dewormed in the last 12 months. Light hookworm infections rarely result in anaemia but may lead to malnutrition, stunting, mental retardation, cognitive and learning deficiencies. The effects of light infection on human health are not immediate and without proper STH programmes consistently implemented in these at risk communities, the effects of STH may impact the future of these infected children. Further, light intensity infection of STH has the potential of sustaining transmission in these endemic areas and could possibly result in heavy infections especially if they are not treated the affected person does not have access to proper nutrition, clean water proper hygiene.

This study further revealed that drinking untreated water directly from the source is one of the risk factors associated with infection STH. An earlier study in Ethiopia also found that 77.5% of households did not treat drinking water and had an increased risk of infection in places with unprotected shallow wells and poor sanitation^32^.

Another significant risk factor identified was keeping untrimmed dirty fingernails. Keeping untrimmed dirty fingernails was taken as a proxy to measure personal hand hygiene. Our findings were in line with a study conducted in South-Central Ethiopia which found that children with untrimmed fingernails were 3.2 times at greater risk of soil-transmitted infections compared to those with trimmed fingernails ^33^. Further, a study in Kenya that assessed multiple locations of soil-transmitted helminth eggs in soil with households in rural setup showed that the highest concentration of eggs in one house occurred in the child’s play area indicating that exposure to household soil would increase the risk of getting infected^34^. This could be a possible explanation for why children in the rural setup of the study, in the younger age group, and with dirty fingernails were more infected because the chances of interacting with infected soils are higher than others in urban areas. Even though another aspect of hand hygiene i.e. handwashing was not found significant in this study, other studies have found that poor or not handwashing was associated with soil-transmitted helminths^35^.

Socio-economic status was not found to be significantly associated with STH infection in this study, although it has been found significant in other studies ^36,37^. Socio-economic status influences access to safe and clean drinking water, adequate sanitation, and improved hygiene facilities and practices.

The public health implications of these findings are that more needs to be done to identify and treat people with STH responsible for sustaining transmission in the communities. This study contributes to the knowledge and understanding of the burden and risk factors of STH in Zambia. Children in schools older than 10 years need to be considered for deworming to reduce the number of reservoirs of parasites and consequently reduce the possibilities of reinfections, especially since the challenge of open defection still exists in these localities^17^. The interventions more than ever need to be more integrated and comprehensive to sustain momentum against this fight to eliminate neglected tropical diseases. Preventive chemotherapy of adolescent girls can for example be integrated into existing health programs like human papillomavirus vaccinations to cover as many girls as possible^15^. Interventions that pertain to health education and behavioral change must be appropriate and comprehensive, targeting all age groups among school-going children creating awareness, improving personal hygiene, ensuring safe and cleaning drinking water, and importance of adequate sanitation among many other important issues. Periodic assessment prevalence, especially in hotspots remains key in tracking progress towards elimination and avoiding neglecting the disease ^2, 15^.

### Study Limitations and Strengths

The study limitations to consider while considering the results are that the answers to the questions were self-reported and therefore guaranteeing accurate information is not possible. The nature of the study design cannot determine the cause and effect^38^. The study used an appropriate study design and correct statistical analysis

## Conclusion and recommendations

Data from this study shows that soil-transmitted helminth infection is present in Katete. The common species was *A. lumbricoides.* The intensity was generally light with a few moderate infections. The factors independently associated with STH infections are drinking untreated water and keeping dirty fingernails. If the prevalence and risk factors associated with soil-transmitted helminths are known, appropriate decision-making based on these data could be expedited as the country races to end/reduce STH infections as a public health problem.

### 5.2 Recommendations

1. The health department working in collaboration with the education department to consider including all school-going children from preschool to grade 12, (where possible not to neglect older children) in the annual deworming to accelerate the reduction of the burden and contribute to the agenda of elimination by 2030.
2. Strengthen behavioral change communication that addresses factors related to personal hygiene and drinking clean and safe water. This can be done in the school health programs by health and education departments
3. Strengthen integrated implementation of interventions in addressing soil-transmitted helminth infections. Leverage existing health programs or platforms to reduce costs. An example is integrating deworming into the Human Papilloma Virus (HPV) vaccination program.

## Supporting information

Ethical approval Malawi IRB

TDRC Approval

NHRA final approval

## Data Availability

All data produced in the present study are available upon reasonable request to the authors

## Abbreviations

AOR: Adjusted Odds Ratio
COMREC: College of Medicine Research Ethics Committee
MDA: Mass Drug Administration
NTD: Neglected Tropical Diseases
OR: Odds Ratio
PSAC: Preschool Children
SAC: School-aged Children
STH: Soil-Transmitted Helminths
WHO: World Health Organization
WRA: Women of Reproductive Age

## Declaration

### Ethical approval and consent to participate

Consent for publication: Not applicable Availability of data and material

Data will be made available from the authors upon request

### Authors contribution

BT: Participated in study design, conducted sample collection, analysis and interpretation. Participated in drafting the manuscript and final approval of the revised manuscript.

WH: Participated in sample collection, sample analysis, interpretation of the results, review of the manuscript and final approval of the revised manuscript. Acted as Guarator

DS: Participated in data analysis, review of the manuscript and final approval of the revised manuscript

ASM: Participated in study design, supervised implementation of the project, interpretation of the results, review of the manuscript and final approval of the revised manuscript.

HH: Participated in study design, supervised implementation of the project, interpretation of the results, review of the manuscript and final approval of the revised manuscript.

## AUTHORS’ INFORMATION AND AFFILIATION

BT (Buumba Tapisha^1,2^) is an Epidemiologist based at Zambart and holds a MSc in Epidemiology.

WH (Westone Hamwata^3^) is a Medical Parasitologist and Entomologist based at the National Health Research and Training Institute and holds a MSc in Medical Parasitology and Master of Public Health.

DS (Danny Sinyange^4^) is a Field Epidemiologist based at The Zambia National Public Health institute and holds a MSc in Epidemiology.

ASM (Adamson S Muula^2^) is a Professor at the University of Malawi, college of Medicine and hold a PhD in Epidemiology

HH (Halwindi Hikabasa^5^; is a Professor at the University of Zambia, School of Health Sciences and holds a PhD in Community Health.

## Acknowledgements

A special thank you to ACEPHEM, Director of Health and District Education Board Secretary in the district the study was conducted for supporting and allowing access to the district laboratory and schools respectively. Special appreciation to the data collectors and data clerk for the data collection.

