## Supplementary material for "Assessment of burden and risk factors associated with Soil-transmitted helminth infections among adolescent girls in Katete District of Zambia: A Cross-Sectional Study": Ethical approval Malawi IRB

### CERTIFICATE OF ETHICS APPROVAL

This is to certify that the College of Medicine Research and Ethics Committee (COMREC) has reviewed and approved a study entitled:

P.06/21/3334 - Assessment of burden and risk factors associated with Soil-transmitted helminth infections among adolescent girls (10-19 years of age) in Katete District of Zambia: A Cross-Sectional Study by Buumba Tapisha

*On 02-Aug-21*

*As you proceed with the implementation of your study, we would like you to adhere to international ethical guidelines, national guidelines and all requirements by COMREC some of which are indicated on the next page for your study*

02-Aug-21

**Prof. E. Umar -Chairperson (COMREC)**

Date

Approved by College of Medicine
02-Aug-2021
(COMREC) Research and Ethics Committee
