## Supplementary material for "Assessment of burden and risk factors associated with Soil-transmitted helminth infections among adolescent girls in Katete District of Zambia: A Cross-Sectional Study": TDRC Approval

**TROPICAL DISEASES RESEARCH CENTRE**
Tel/Fax +260212 615444

P O Box 71769
Ndola,
ZAMBIA

**TDRC ETHICS REVIEW COMMITTEE**
**IRB REGISTRATION NUMBER : 00002911**
**FWA NUMBER : 00003729**

TRC/C4/05/2021

21st May 2021

Buumba Tapisha
College of Medicine
University of Malawi
Blantyre, Malawi

Dear Mr Buumba

**RE: ETHICAL APPROVAL OF STUDY PROTOCOL**

Reference is made to the protocol entitled **"Assessment of burden and risk factors associated with Soil transmitted helminth infections among adolescent girls (10-19 years of age) in Katete District of Zambia: A Cross-Sectional Study",** which was submitted to the TDRC Ethics Review Committee for review.

On behalf of the Chairperson of the Committee, I wish to inform you that the Committee reviewed and approved your study protocol.

You are further required to submit progress reports to the TDRC ERC twice a year.

Should there be any protocol modifications or amendments, you are required to notify the ERC and submit protocol amendments for approval.

You are now required to submit your protocol to the National Health Research Authority for final approval following the link: https://www.nhra.org.zm. A final report of the study should be submitted to the Ethics Review Committee Secretariat at the end of the study.

**This approval is valid for the period 21st May 2021 to 20th May 2022.**

The Committee wishes you success in the execution of the study.

Yours faithfully,
**TROPICAL DISEASES RESEARCH CENTRE**

Sydney Mwanza
**DEPUTY SECRETARY – TDRC Ethics Review Committee**

TROPICAL DISEASES RESEARCH CENTRE
ETHICS REVIEW COMMITTEE
21 MAY 2021
P.O. BOX 71769
NDOLA, ZAMBIA
