## Supplementary material for "Assessment of burden and risk factors associated with Soil-transmitted helminth infections among adolescent girls in Katete District of Zambia: A Cross-Sectional Study": NHRA final approval

### NATIONAL HEALTH RESEARCH AUTHORITY

**Paediatric Centre of Excellence, University Teaching Hospital, P.O. Box 30075, LUSAKA**

Tell: +260211 250309 | | www.nhra.org.zm


**Ref No: NHRA000013/31/05/2021** **Date: 31st May, 2021**

The Principal Investigator,
Buumba Tapisha
College of Medicine,
University of Malawi,
P/Bag 360,
Chichiri, Blantyre 3,
Malawi**.**

Dear Buumba Tapisha,

### Re: Request for Authority to Conduct Research

The National Health Research Authority is in receipt of your request for authority to conduct research titled **"ASSESSMENT OF BURDEN AND RISK FACTORS ASSOCIATED WITH SOIL-TRANSMITTED HELMINTH INFECTIONS AMONG ADOLESCENT GIRLS (10-19YEARS OF AGE) IN KATETE DISTRICT OF ZAMBIA: A CROSS-SECTIONAL STUDY."** I wish to inform you that following submission of your request to the Authority, our review of the same and in view of the ethical clearance, this study has been **approved** on condition that:

1. The relevant Provincial and District Medical Officers where the study is being conducted are fully appraised;
2. Progress updates are provided to NHRA quarterly from the date of commencement of the study;
3. The final study report is cleared by the NHRA before any publication or dissemination within or outside the country;
4. After clearance for publication or dissemination by the NHRA, the final study report is shared with all relevant Provincial and District Directors of Health where the study was being conducted, University leadership, and all key respondents.

Yours sincerely,

Prof. Godfrey Biemba
Director/CEO
**National Health Research Authority**
